# A low-cost, time-efficient, sensitive quantitative thin layer chromatography reveals unaltered exogenous sphingosine utilisation from erythrocytes of MAFLD patients

**DOI:** 10.64898/2026.07.04.26357124

**Authors:** Eleftheria Spourita, Konstantinos Mimidis, Ioannis Tentes, Konstantinos Anagnostopoulos, Charalampos Papadopoulos

**Author notes:** Address correspondence to this author. Laboratory of Biochemistry, Department of Medicine, Democritus University of Thrace, Dragana, University Campus, Alexandroupolis, Greece, 68100.

## Abstract

**BACKGROUND:** Erythrophagocytosis constitutes a major pathogenic mechanism of metabolic dysfunction associated fatty liver disease (MAFLD). Our previous research established a quantitative thin-layer chromatography (TLC) technique for sphingomyelin, revealing reduced levels in the red blood cells (erythrocytes) of patients with metabolic dysfunction associated fatty liver disease (MAFLD). This reduction was accompanied by erythrocyte sphingosine accumulation, a driver of pro-inflammatory erythrophagocytosis, though sphingosine 1-phosphate release remained stable. To better understand erythrocyte sphingosine metabolism, we adapted our quantitative TLC method to analyze sphingosine within the erythrocyte-conditioned media (ECM) of MAFLD patients.

**Methodology:** Separation was performed on 10X10cm Silica gel 60 F254 plates using a mobile phase of chloroform, methanol, acetic acid, and water (60:50:1:4 v/v/v/v). The dynamic range, linearity, and range of linearity were assessed by analysing sphingosine levels from 0.1 to 100μg/spot. We validated the system’s precision and sensitivity by performing triplicate analyses of sphingosine standards (1.25, 2.5, and 5μg). The limits of detection and quantification were derived from the calibration curve’s slope and standard deviation (3.3 XSD/slope for LOD; 10 XSD/slope for LOQ). Accuracy was assessed via recovery tests at 100%, 200%, and 300% of a 2.5μg load. We confirmed specificity by evaluating the retention factors against other lipid species. This protocol was applied to Folch-extracted lipids from the ECM (5 × 10^7^ cells/ml) of four MAFLD patients and four healthy controls, spiked with 5μg of sphingosine.

**Findings:** The calibration model, based on combined Green and Blue color intensities, followed the linear equation y = -11.171x + 353.25(R^2^ = 0.94). Interday precision values were 0.21%, 1.65%, and 0.44%, while recovery rates (accuracy) ranged from 94.5% to 98.7%. The measured LOD and LOQ were 0.75μg and 1.21μg, respectively. The sensitivity was calculated at 90ng. Statistical analysis showed no significant variance in sphingosine concentrations in erythrocyte-conditioned media between the MAFLD group and the control group.

**Summary:** The described thin layer chromatography is accurate, precise, sensitive, with good limits of detection and quantification, and most importantly is low-cost and time-efficient. Using this method, we show that while erythrocytes of MAFLD patients exhibit sphingosine accumulation, the utilisation of exogenous sphingosine from their erythrocytes is not affected. This suggests that the metabolic shift may be driven by increased sphingosine supply from the plasma.

## INTRODUCTION

Erythrocyte removal represents a major pathogenic mechanism in metabolism dysfunction associated fatty liver disease (MAFLD)(1). Previous studies have shown that erythrocytes of MAFLD patients exhibit various pro-phagocytic signals, such as augmented monocyte chemoattractant protein 1 (MCP1)(2,3), reduced cluster of differentiation 47 (CD47)(3), increased membrane sphingosine, but reduced opsonization of exposed phosphatidylserine(4). Interestingly, erythrocyte sphingosine accumulation has been found to function as an additional phagocytosis signal, independent of phosphatidylserine exposure or CD47 reduction(5). Importantly, sphingosine-mediated erythrocyte removal is pro-inflammatory(5), possibly due to the effect on intracellular organelle membrane integrity of the phagocyte(6).

The molecular determinants of erythrocyte sphingosine accumulation in MAFLD have not been clarified yet. Accordingly, we showed previously that erythrocytes of MAFLD patients exhibit unaltered capacity of sphingosine 1-phosphate (S1P) release(2). This shows that the mechanisms affecting sphingosine 1-phosphate release are intact on erythrocytes of MAFLD patients. Hence, erythrocyte sphingosine accumulation is not driven by Mfsd2b, BAND3, or ABCA1 dysfunction(7,8,9). Another explanation could be the reduced activity of sphingosine kinase(10). Relatively, in our previous study, we reported that in vitro erythrocyte sphingosine accumulation can be driven by inhibition of sphingosine kinase(3). Because sphingosine kinase activity positively regulates exogenous sphingosine uptake and utilisation, we hypothesized that if sphingosine kinase is inhibited on erythrocytes of MAFLD patients, the sphingosine content of erythrocyte-conditioned media of MAFLD patients would be increased. Otherwise, erythrocyte sphingosine accumulation in MAFLD would be triggered by augmented sphingosine supply(11), i.e., from plasma, since de novo sphingosine synthesis or ceramide-derived sphingosine formation are not active in human erythrocytes (10). Thus, we decided to measure erythrocyte-conditioned media sphingosine levels from MAFLD patients and healthy controls.

The measurement of sphingosine can be executed by various new technologies.

However, thin layer chromatography remains a cost-effective, time-efficient, precise, and accurate method. We previously developed a method for measuring sphingomyelin from erythrocytes(12). Based on that methodology, we previously developed and validated a method for sphingosine, without describing the detailed process for developing and validating that method. In this study, we describe the validation and application of this method.

## METHODS

### 2.1 MAFLD patients and healthy volunteers

Four patients with MAFLD (3 men, 1 woman, aged 47.3±2 years) and 4 healthy controls (2 men, 2 women, aged 39,3±5,8 years) participated in our study and were recruited by the 1st Internal Medicine Clinic of the University Hospital of Alexandroupolis. MAFLD was verified by ultrasound and blood work. The Scientific Council of the University Hospital of Alexandroupolis and the Ethics Committee approved the study after obtaining each participant’s informed consent.

### 2.2 Isolation of red blood cells

A blood sample (3 ml) containing EDTA was centrifuged at 200 g for 10 minutes at 4°C. Plasma and white blood cells were removed. Then, the red blood cell pellet was washed with cold serum and centrifuged at 200 g for 10 minutes at 4°C. This step was repeated a total of 4 times.

### 2.3 Isolation of red blood cell membranes

Red blood cells that had been isolated as previously were dissolved (1:10) with hemolysis buffer (Tris 1mM, NaCl 10mM, EDTA 1mM, pH 7.2) and incubated at 4°C for 30 minutes with continuous shaking. Then, they were centrifuged at 15,000 rpm at 4°C for 15 minutes. The precipitate was collected, and the previous step was repeated until the precipitate turned milky white, indicating the removal of hemoglobin. The samples were stored at -80°C.

### 2.4 Production of erythrocyte-conditioned media

Red blood cells (5*10^9^/ml) were incubated in RPMI 1640 containing 10% FBS (v/v), 1%(v/v) streptomycin/penicillin, in 5% CO2, at 37°C for 24 hours. Then, the erythrocyte-conditioned medium of both patients and healthy volunteers was collected by centrifugation at 200 g for 10 minutes. The samples were stored at -80°C. As a control, culture medium was placed in parallel in 35mm diameter dishes, under the same conditions, and the same procedure was followed. To exclude hemolysis, a spectrophotometer was used, and no hemoglobin was detected in the culture medium from the red blood cells.

### 2.5 Lipid isolation

500 μL of conditioned erythrocyte medium was used. For all samples, the Folch method (13) was used, i.e., chloroform/methanol (2:1). The solvent/sample mixture ratio was 1:1. The mixture was centrifuged at 2000 rpm for 5 minutes. Then, the organic phase was collected in a different tube, and 500 μL of the solvent mixture was added to the polar phase. Centrifugation was performed at 2000 rpm for 5 minutes, and the organic phase was collected. Then, water was added to the organic phase with an organic phase/water ratio (1:1), and centrifugation was performed at 2000 rpm for 5 minutes. Finally, the tubes were placed at 80° C until the solvent evaporated.

### 2.6 Lipid separation and imaging

The thin layer chromatography analysis was performed on a 10X10 cm plate (silica gel 60 F254 Merck KGaA 64071 Darmstadt, Germany). A chloroform/methanol/acetic acid/water mixture (60/50/1/4) (v/v/v/v) was used as the mobile phase. Initially, the plate was placed at 150° C for 10 minutes and developed in the mobile phase solvent mixture. Subsequently, the samples were loaded and developed in a suitable closed container with the mobile phase. After lipid separation, the plate was dried with hot air and placed in a closed container containing 3.5 g iodine for 40 minutes at room temperature. Lipid bands appeared with a dark yellow color. The lipids identified based on appropriate controls were cholesterol, phosphatidylethanolamine (PE), sphingosine, phosphatidylinositol, phosphatidylcholine, and sphingomyelin.

### 2.7 Thin Layer Chromatography Development

The TLC chromatograms were digitized with a Canon MX-475 scanner in color format at a resolution of 300 dpi. The determination of the intensity of the lipid bands was done using the Microsoft Paint program, and with the ‘pick color’ tool, the intensity of the green and blue colors was measured from the middle of the band (14).

### 2.8 Linearity of TLC

To determine the linearity, analysis was performed at 9 different concentrations (100, 50, 25, 10, 5, 2.5, 1.25, 0.75, 0.5 μg/point) and the experiments were repeated three times on different days.

### 2.9 PRECISION OF TLC

Repeatability was expressed as relative standard deviation (coefficient of variation, CV%) for 1.25, 2.5, and 5 μg/spot

### 2.10 ACCURACY OF TLC

To study the accuracy, the recovery rate was studied for 100%, 200%, and 300% of the mass added at 2.5 μg/point.

### 2.11 LIMITS OF DETECTION AND QUANTIFICATION

The limit of detection (LOD) and the limit of quantification (LOQ) were determined after experiments repeated three times for the three concentrations (0.75, 0.5, 0.25 μg/point). The LOD and LOQ are expressed as 3.3XSD/slope of the repeatability and 10XSD/slope of the repeatability, respectively.

### 2.12 SENSITIVITY OF TLC

The sensitivity was expressed as the ratio of the slope that described the linearity of the method to 1unit of mass change (1μg/spot).

### 2.13 SPECIFICITY OF TLC

The specificity of the tlc was examined by comparing the retention factors of various lipids using the same stable and mobile phase of our system.

### 2.14 Statistical Analysis

Sphingosine levels (μg/spot after adjustment for a 5 μg spike, TLC□derived) were compared between Patients□ECM and Healthy□ECM using a Bayesian independent□samples t-test. This approach was used because it does not require large samples and has the ability to give reliable results in small sample sizes.

The primary estimand was the difference in group means (Patients − Healthy) in raw units (μg/spot). We adopted the Jeffreys–Zellner–Siow (JZS) prior on the standardized effect size 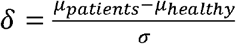, i.e., a *Cauchy*(0, 0.707) prior (default “medium” scale), with default objective priors on the grand mean and the common standard deviation, as implemented in the BayesFactor package (R). Posterior inference was obtained by sampling from the posterior distribution using posterior(); we report posterior means with **95% highest**□**density credible intervals (HDIs)** for both the raw difference and the standardized effect (δ). Evidence for a non□zero difference was quantified with the Bayes factor *BF*_lO_ (alternative vs. point null). All analyses were performed in R.

## RESULTS

### 3.1 Linearity of thin layer chromatography

Thin Layer Chromatography analysis was performed for sphingosine at concentrations (100, 50, 25, 10, 5, 2.5, 1.25, 0.75μg/point) (figure 14) and the colors Red, Green, Blue, Red + Green, Red + Blue, Green + Blue were analyzed. The results are presented in detail in Table 1. Then, the average of the results was calculated.

**Table 1:** Performance characteristics of the quantitative thin layer chromatography for sphingosine.

| Performance characteristic | Value |
| --- | --- |
| Linearity ($\mu\text{g/spot}$) | 1.25-5 |
| Limit of detection ($\mu\text{g/spot}$) | 0.75 |
| Limit of quantification ($\mu\text{g/spot}$) | 1.21 |
| Range ($\mu\text{g/spot}$) | 1-100 |
| Inter-day CV (%) (2.5$\mu\text{g}$) | 0.21 |
| Inter-day CV (%) (5$\mu\text{g}$) | 1.65 |
| Inter-day CV (%) (10$\mu\text{g}$) | 0.44 |
| Recovery (%) (+100%) | 97.0±1.6 |
| Recovery (%) (+200%) | 98.7±8.4 |
| Recovery (%) (+300%) | 94.5±8.8 |
| Sensitivity | 90ng |
| Specificity (retention factor) | 0.32 |

The thin layer chromatography method for sphingosine shows linearity over a range of concentrations (1.25-5μg/point), based on the composition of the red, green, and blue colors, as well as their combination. The best linearity is shown by the combination of the colors green and blue (G+B), with the linearity equation being y = -11.171x+353.25, with R^2^=0.94, as shown in Figure 1.

**Figure 1:**
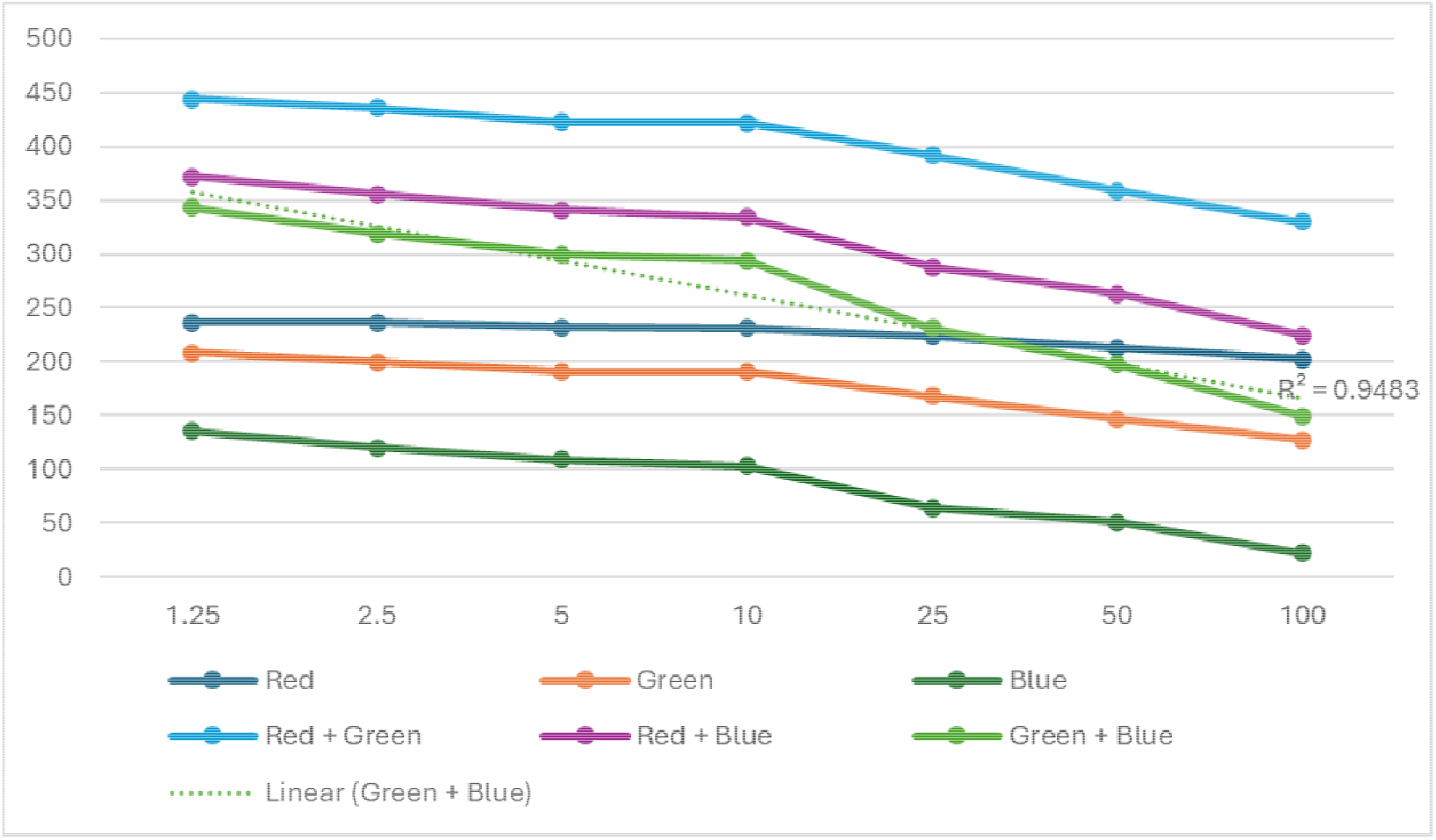
Linearity of the quantitative thin layer chromatography for sphingosine using different combinations of color intensities.

### 3.2 Dynamic range, specificity, sensitivity, accuracy, and precision of quantitative thin layer chromatography

Next, the dynamic range, detection limit, and quantitation limit, as well as the repeatability and precision, expressed as %, of the method were examined. The dynamic range was determined to be 1-100 μg/point (Figure 2).

**Figure 2:**
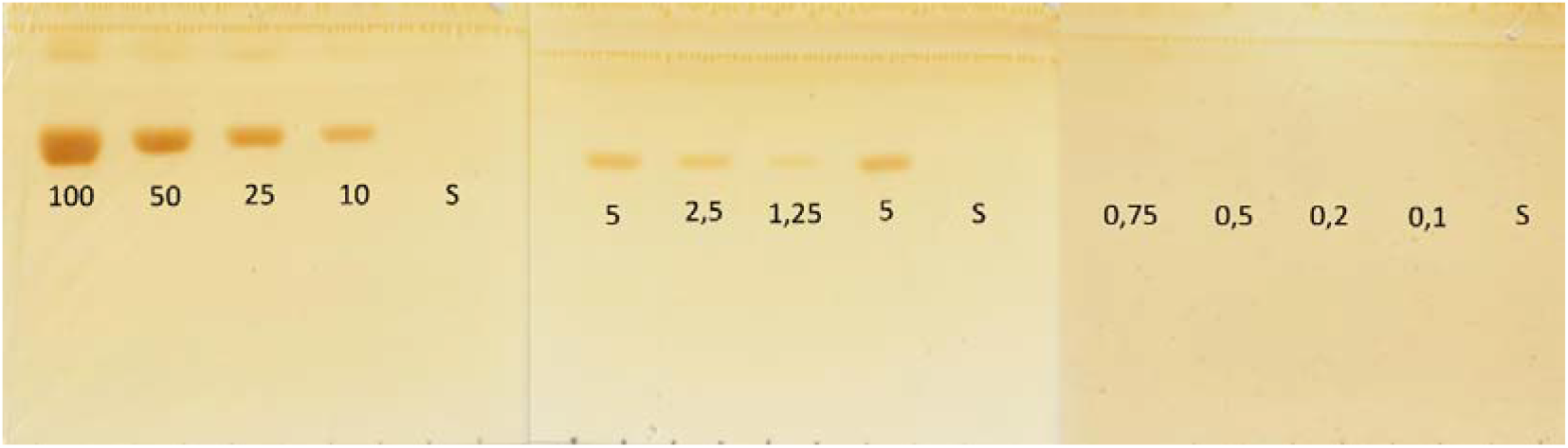
Dynamic range of the quantitative thin layer chromatography for sphingosine

The detection limit is 0.75 μg/point, while the quantification limit is 1.21 mg/point. The repeatability is 0.21%, 1.65%, and 0.44% for the masses of 2.5μg, 5μg, and 10μg, respectively. Finally, the precision was expressed as % recovery and for this method it is 97.0±1.6%, 98.7±8.4%, and 94.5±8.8% for 2.5 + 100%, 200%, and 300% respectively (Figure 2), while the sensitivity of the method was calculated at 11.1, and was further validated at concentration 5, 5.1, 5.2 μg(Figure 3) The characteristics of the TLC method for sphingosine are summarized in Table 1.

**Figure 3:**
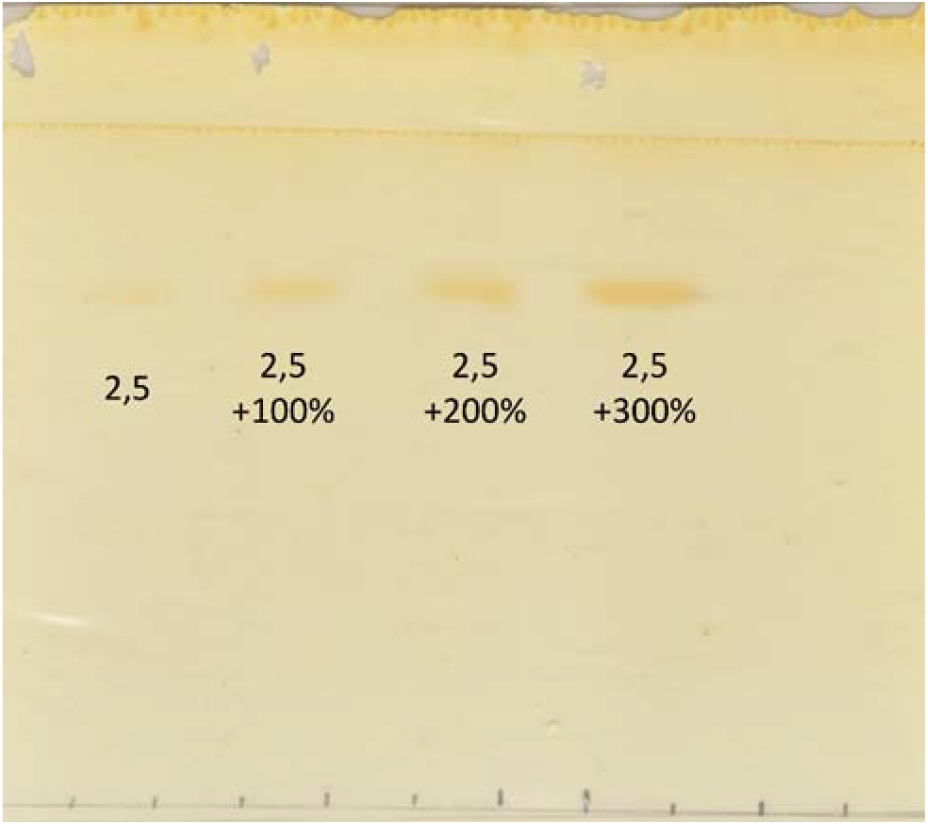
Precision of the quantitative thin layer chromatography for sphingosine

**Figure 4:**
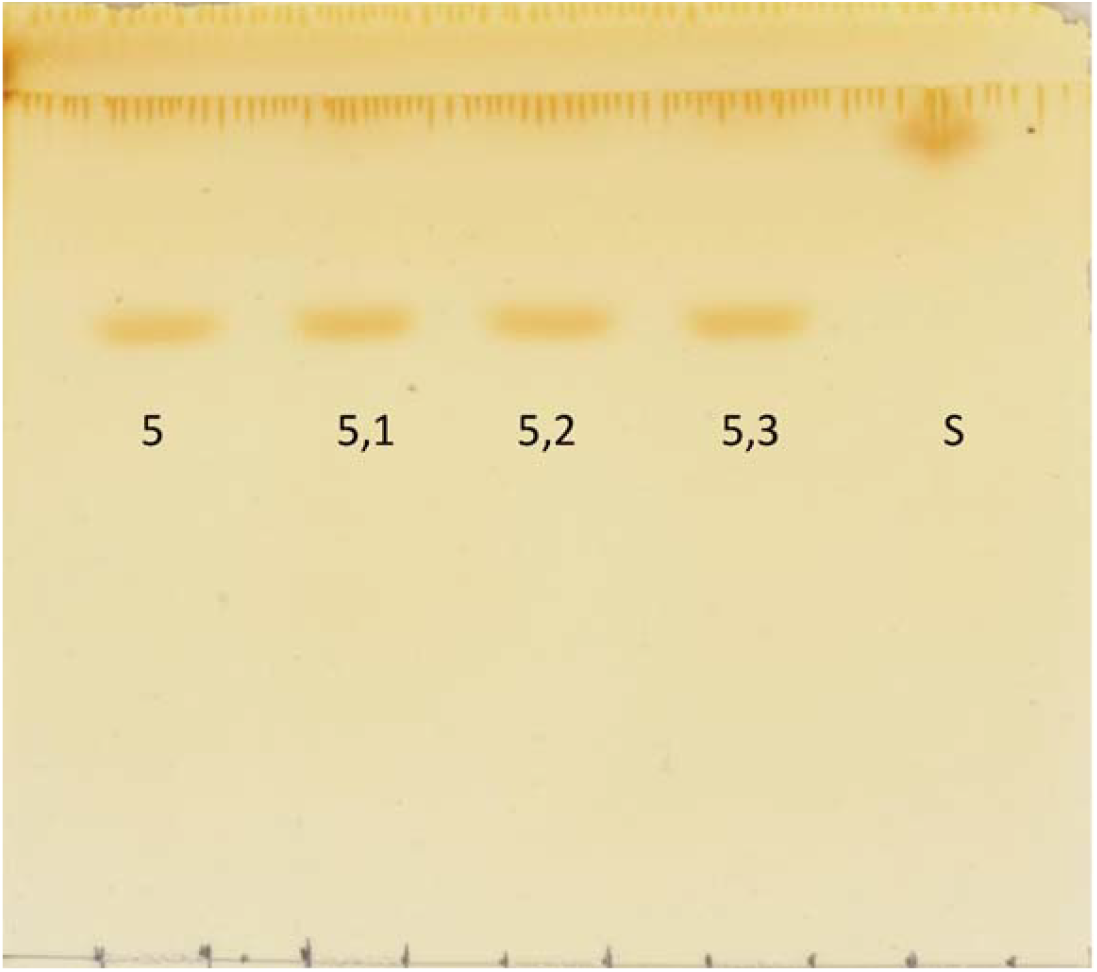
Validation of the sensitivity of the quantitative thin layer chromatography for sphingosine

The specificity of the method was found to be sufficient, assuming a longer distance for the development of the mobile phase (10cm). Sphingosine exhibits a separate retention factor from free cholesterol, phosphatidylethanolamine, phosphatidylserine, phosphatidylinositol, phosphatidylcholine, sphingomyelin, and nonpolar lipids (esterified cholesterol or triglycerides) (Table 2). However, the retention factor of sphingosine is close enough to that of phosphatidylethanolamine to require at least 10cm of distance for the mobile phase.

**Table 2:** Specificity of the quantitative thin layer chromatography for sphingosine. *FChol: free* cholesterol; PC: phosphatidylcholine; PE: phosphatidylethanolamine; PI: phosphatidylinositol; PSer: phosphatidylserine; SM: sphingomyelin; Sph: sphingosine.

|  | FChol | PE | Sph | PSer | PI | PC | SM |
| --- | --- | --- | --- | --- | --- | --- | --- |
| R<sub>f</sub> | 0.053 | 0.282 | 0.35 | 0.443 | 0.594 | 0.75 | 0.863 |

### 3.3 Application of quantitative thin layer chromatography for the analysis of sphingosine in erythrocyte-conditioned media from patients with MAFLD and from healthy volunteers

4 patients and 4 healthy volunteers participated in the study. The amount of Sphingosine in 500μL conditioned erythrocyte media was measured and compared between patients with MAFLD and healthy volunteers. In order to measure the amount of sphingosine, as it is less than the quantitation limit of the method in the biological sample, 5μg of pure sphingosine was added to each sample. No statistically significant difference was observed in the amount of sphingosine in the conditioned media of erythrocytes of patients with MAFLD (93.8%±19,4% of growth medium alone) compared to healthy volunteers (of growth medium alone) table 3.

**Table 3:** Application of the quantitative thin layer chromatography for sphingosine on erythrocyte -conditioned media of MAFLD patients and healthy controls. Results expressed as a percentage of the sphingosine content in the growth medium alone. *MAFLD: metabolic dysfunction associated fatty liver disease*.

| HEALTHY CONTROL S (n=4) | MAFLD PATIENTS (n=4) | GROWTH MEDIUM(n=2) |
| --- | --- | --- |
| 76.9%±39.6 % | 93.8%±19,4 % | 100,00%±16,8% |

#### Patients-ECM vs Healthy-ECM

Sphingosine levels (μg/spot) were slightly higher in Patients-ECM (mean 2.252, SD 0.611, *n*=4) than in Healthy-ECM (mean 2.095, SD 0.865, *n*=4), yielding an observed difference of **+0.157** μ**g/spot**. Under a Bayesian independent two-sample normal model with a common variance and an objective prior p(*µ*_*Patients*_, *µ*_*Healthy*_, *σ*) ∝ 1/*σ*, the **posterior mean difference** (Patients − Healthy) was **0.157** μ**g/spot** with a **95% credible interval** of **[−1.143, 1.453]** μ**g/spot** and **Pr(**Δ**>0) = 0.61**. The **standardized effect** had a posterior mean of **0.20** with a **95% credible interval [−1.19, 1.59]**. Using a **region of practical equivalence (ROPE)** on the raw difference defined a priori as ±0.2 × *s*_*pooled*_ = ±0.150μg/spot, **20.2%** of the posterior mass lay within the ROPE (10.2% for ±0.1 × *s*_*pooled*_). Complementarily, a JZS Bayes-factor *t*-test (Cauchy prior on awith r = 0.707) yielded **BF**□□ **= 0.54** (i.e., **BF**□□ **= 1.86**), indicating **weak evidence** for no difference; sensitivity analyses at *r* = 1.0and *r* = 1.414gave **BF**□□ **= 0.44** and **0.35**, respectively.

## DISCUSSION

Metabolic dysfunction associated fatty liver disease is characterized by increased hepatic erythrocyte removal(1). Although in animal models, erythrocyte phosphatidylserine exposure and its recognition by opsonins drive erythrophagocytosis(1,15), in humans, the exact mechanisms have not been elucidated yet. We previously showed reduced phosphatidylserine opsonization of erythrocytes in human MAFLD(4). Instead, we showed that erythrocytes of MAFLD patients exhibit reduced CD47(3), increased MCP1 binding(3) and release(2), and sphingosine accumulation(3). Erythrocyte sphingosine accumulation is an important determinant of erythrocyte removal that is independent of other signals(5). More importantly, sphingosine renders erythrophagocytosis pro-inflammatory(5). We thus decided to explore further the molecular mechanisms for the erythrocyte sphingosine accumulation in MAFLD.

Erythrocyte sphingosine accumulation can be triggered by reduced sphingosine kinase activity(10). Indeed, we showed that in vitro this could be a rational explanation(3). However, if this were the case for MALFD, then exogenous sphingosine utilisation from erythrocytes of MAFLD patients would be decreased. For this reason, we examined the content of sphingosine in erythrocyte-conditioned media of MAFLD and healthy controls.

To study sphingosine, we developed and validated a quantitative thin layer chromatography method based on our previous study for the quantitation of sphingomyelin(12). Our method is linear over a wide range of mass, accurate, precise, sensitive, and exhibits low limits of detection and quantification. It is, however, more crucial that our method is low-cost and time-efficient. This method only requires the separation of lipids on a silica plate, developed with a mixture of chloroform, methanol, acetate, and water, visualization with iodine, and measurement of the intensity of the blue and green color. We should, however, mention that our method has some limitations, namely: the limit of quantification (1.21μg), sensitivity (90ng), and the retention factor that is close to that of phosphatidylethanolamine. The problem of the limit of quantification can be overcome through the use of a spike, i.e., the pre-load with a standard of known mass. The limitation of the sensitivity can be overcome through the use of higher volumes of sample used for lipid separation. Finally, the drawback of the retention factor can be easily overcome through the use of a longer distance of development for the mobile phase.

Using this quantitative thin layer chromatography, we report unaltered exogenous sphingosine utilisation from erythrocytes of MAFLD patients. This shows that erythrocyte sphingosine accumulation in MAFLD is not driven by inhibition of sphingosine kinase activity, but possibly by augmented supply of sphingosine from plasma in vivo. Other explanations, such as reduced efflux of sphingosine 1-phosphate, can be rejected based on our previous study of unaltered S1P in erythrocyte conditioned media from MAFLD patients and healthy controls(2). In addition, the production of sphingosine either through de novo synthesis or through ceramide hydrolysis has been shown to be minimal(10).

Our study has some limitations. Firstly, our method can be used to assess only clinically significant differences. The sensitivity of our method doesn’t allow the detection of smaller differences. Secondly, our method requires the use of a spike to measure samples that are below the limit of quantification. Finally, the number of patients we used was limited. Our adapted robust statistical analysis revealed weak evidence for no alteration in the sphingosine utilisation from erythrocytes of MAFLD patients compared to the healthy controls. For these reasons, our results should be interpreted with caution.

Despite the limitations of our study, our results certainly shed light on the metabolism of erythrocyte sphingosine in human metabolic inflammation of the liver. More importantly, our method provides a means of sphingosine quantitation for laboratories with a low budget and/or time.

## CONCLUSIONS

In conclusion, thin layer chromatography-based separation of lipids, coupled with iodine staining and green and blue color intensity measurement, is a low -cost, time – efficient, and high-performance method for sphingosine. We reveal that the sphingosine -dependent pro-inflammatory erythrophagocytosis in MAFLD is not driven by augmented exogenous sphingosine utilisation from erythrocytes.

## Data Availability

All data produced in the present study are available upon reasonable request to the authors

## ABBREVIATIONS

Cd47: cluster of differentiation 47
MAFLD: metabolism dysfunction associated fatty liver disease
MCP1: monocyte chemoattractant protein 1
FChol: free cholesterol
PC: phosphatidylcholine
PE: phosphatidylethanolamine
PI: phosphatidylinositol
PSer: phosphatidylserine
SM: sphingomyelin
SPh: Sphingosine

## Authors’ contribution

CP designed the research/study; ES, KM, IT, and CP performed the research/study; ES and CP collected data; CP and KA analyzed data; CP wrote the manuscript; and CP and KA supervised the study.

